# Liver injury is associated with severe Coronavirus disease 2019 (COVID-19) infection: a systematic review and meta-analysis of retrospective studies

**DOI:** 10.1101/2020.04.09.20056242

**Authors:** Mohammad Parohan, Sajad Yaghoubi, Asal Seraj

## Abstract

**Aim:** The Coronavirus disease 2019 (COVID-19) outbreak is a major threat to human beings. Lung injury has been reported as the major outcome of COVID-19 infection. However, liver damage has also been considered to occur in severe cases. Current meta-analysis of retrospective studies was done to summarize available findings on the association between liver injury and severity of COVID-19 infection.

**Methods:** Online databases including PubMed, Scopus, Web of Science and Cochrane Library were searched to detect relevant publications up to 1 April 2020, using relevant keywords. To pool data, a fixed- or random-effects model was used depending on the heterogeneity between studies. Furthermore, publication bias test and sensitivity analysis were also done.

**Results:** In total, 20 retrospective studies with 3,428 COVID-19 infected patients (severe cases = 1,455 and mild cases = 1,973), were included in this meta-analysis. Higher serum levels of Aspartate aminotransferase (weighted mean difference = 8.84 U/L, 95% CI = 5.97 to 11.71, P<0.001), Alanine aminotransferase (weighted mean difference = 7.35 U/L, 95% CI = 4.77 to 9.93, P<0.001), total Bilirubin (weighted mean difference = 2.30 mmol/L, 95% CI = 1.24 to 3.36, P<0.001) and lower serum levels of Albumin (weighted mean difference = −4.24 g/L, 95% CI = −6.20 to −2.28, P<0.001), were associated with a significant increase in the severity of COVID-19 infection.

**Conclusions:** The incidence of liver injury seems to be higher in patients with severe COVID-19 infection. This finding could help physicians to identify patients with poor prognosis at an early stage.

## Introduction

In December 2019, a cluster of severe acute respiratory syndrome (SARS), now known as Coronavirus disease 2019 (COVID-19), occurred in Wuhan city, the capital of Hubei Province, China ^1-3^. The disease has rapidly spread from China to other countries. As of April 4, 2020, a total of 1,051,635 COVID-19 confirmed cases and 56,985 deaths in 206 countries and territories have been reported ^4^. Full-genome sequencing indicated that COVID-19 is a distinct clade from the beta-coronaviruses associated with human SARS and Middle East respiratory syndrome (MERS) ^5^.

SARS, MERS and COVID-19 can cause intestinal, respiratory, neuronal and hepatic diseases, and may lead to respiratory distress syndrome, organ failure, and even death in severe cases ^5-7^. Several studies have reported the clinical characteristics and laboratory findings associated with different degrees of liver injury in patients with COVID-19 infection ^8-27^. We are aware of no meta-analysis that summarized available findings in this regard. Thus, in this systematic review and meta-analysis, the laboratory findings and mechanism of liver injury caused by COVID-19 infection were summarized.

## Materials and methods

### Study protocol

A systematic literature search and a quantitative meta-analysis were planned, conducted and reported according to the Preferred Reporting Items for Systematic Reviews and Meta-Analyses (PRISMA) guidelines ^28^.

### Search strategy

We conducted a literature search using the online databases of PubMed, Scopus, Web of Science and Cochrane Library for relevant publications up to 1 April 2020. The following medical subject headings (MeSH) and non-MeSH keywords were used in our search strategy: (“COVID-19” OR “severe acute respiratory syndrome coronavirus 2” OR “SARS-CoV-2” OR “novel coronavirus” OR “2019-nCoV”) AND (“Alanine Transaminase” OR “Alanine aminotransferase” OR “SGPT” OR “Aspartate Aminotransferases” OR “SGOT” OR “Bilirubin” OR “Serum Albumin” OR “Liver”). The literature search was performed by two reviewers (MP and SY). We also searched the reference lists of the articles to identify missed studies. No restriction was applied on time of publication and language. To facilitate the screening process of studies from online databases, all search results were downloaded into an EndNote library (version X8, Thomson Reuters, Philadelphia, USA). The search strategy is presented in detail in Supplementary Table 1.

### Eligibility Criteria

Studies were included if they met the following inclusion criteria: (1) observational studies with retrospective design; (2) all articles assessing the association between serum levels of Aspartate aminotransferase (AST), Alanine aminotransferase (ALT), Albumin, Bilirubin and severe outcome from COVID-19 infection as the major outcomes of interest and reported mean (SD) or median (IQR) for serum levels of AST, ALT, Albumin, Bilirubin in both severe and non-severe COVID-19 infected patients. Expert opinion articles, review articles, books and theses were excluded.

### Data extraction and assessment for study quality

Two reviewers (MP and AS) extracted the following data from the studies: author’s name, publication year, study design, sample size, age and gender of patients, serum levels of AST, ALT, Albumin and Bilirubin and outcome assessment methods.

The Newcastle–Ottawa Scale was used for assessing the quality of the included studies ^29^. Based on the NOS, a maximum of nine points can be awarded to each article. In this review, studies with the Newcastle–Ottawa Scale score of ≥ 5 were considered as high quality publications.

### Statistical analysis

Mean (SD) or median (IQR) for serum levels of AST, ALT, Albumin and Bilirubin were used to estimate the effect size. The fixed or random-effect model was used based on heterogeneity test. The heterogeneity between studies was evaluated using the Cochrane Q test ^30^. The publication bias was evaluated by the visual inspection of funnel plot and Egger’s regression tests ^31^.The sensitivity analysis was done to assess the effect of each study on the pooled effect size. All statistical analyses were performed using the Stata 14 software package (Stata Corp, College Station, TX, USA).

## Results

### Search results

Overall, 212 articles were identified in our initial literature search. Of these, 35 duplicates, 29 non-English, 3 non-human, 18 reviews and 95 papers that did not fulfill our inclusion criteria were excluded, leaving 32 articles for further evaluation. Out of remaining 32 articles, 12 were excluded because of the following reason: did not report mean (SD) or median (IQR). Finally, we included 20 articles in this systematic review and meta-analysis (Figure 1).

**Figure 1.**
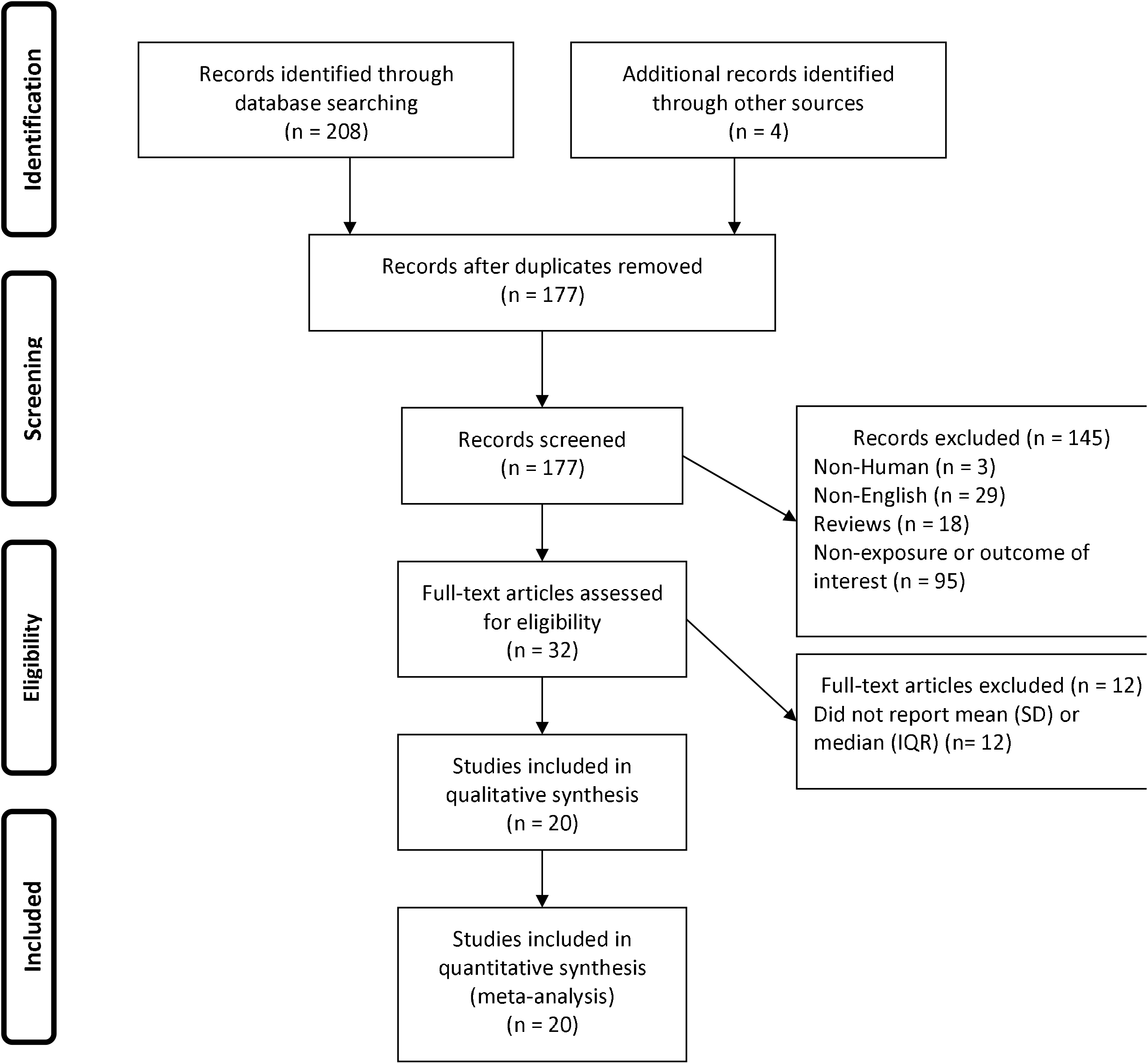
Flow chart of study selection.

### Study characteristics

All studies were conducted in China and used retrospective design ^8-27^. The sample size of studies ranged from 21 to 651 patients (mean age, 53.3 years). All studies used real-time reverse transcriptase–polymerase chain reaction (RT-PCR) to identify COVID-19 infection. The Newcastle–Ottawa Scale scores ranged between 4 to 9. The characteristics of the included articles are presented in Table 1.

**Table 1.**
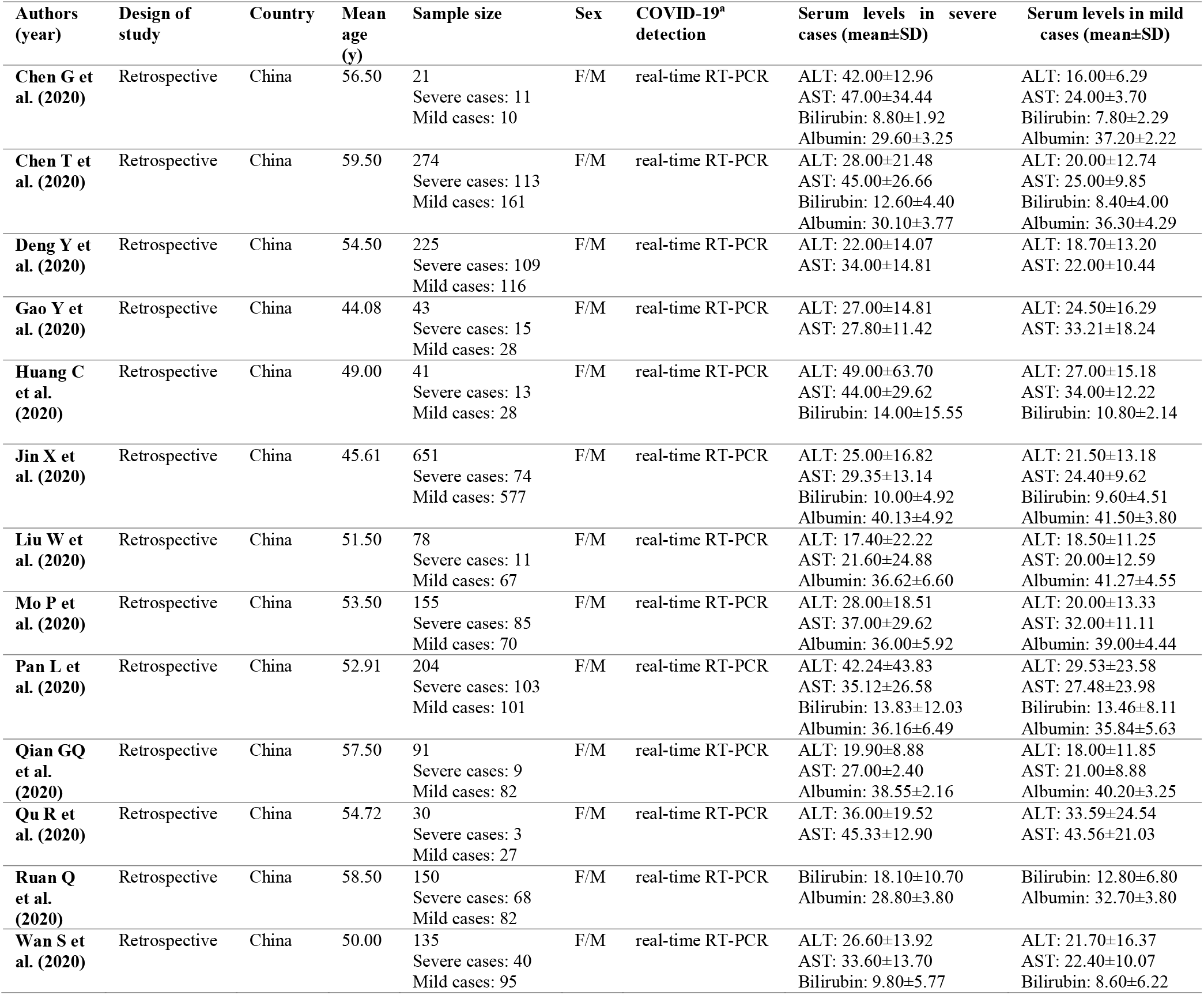
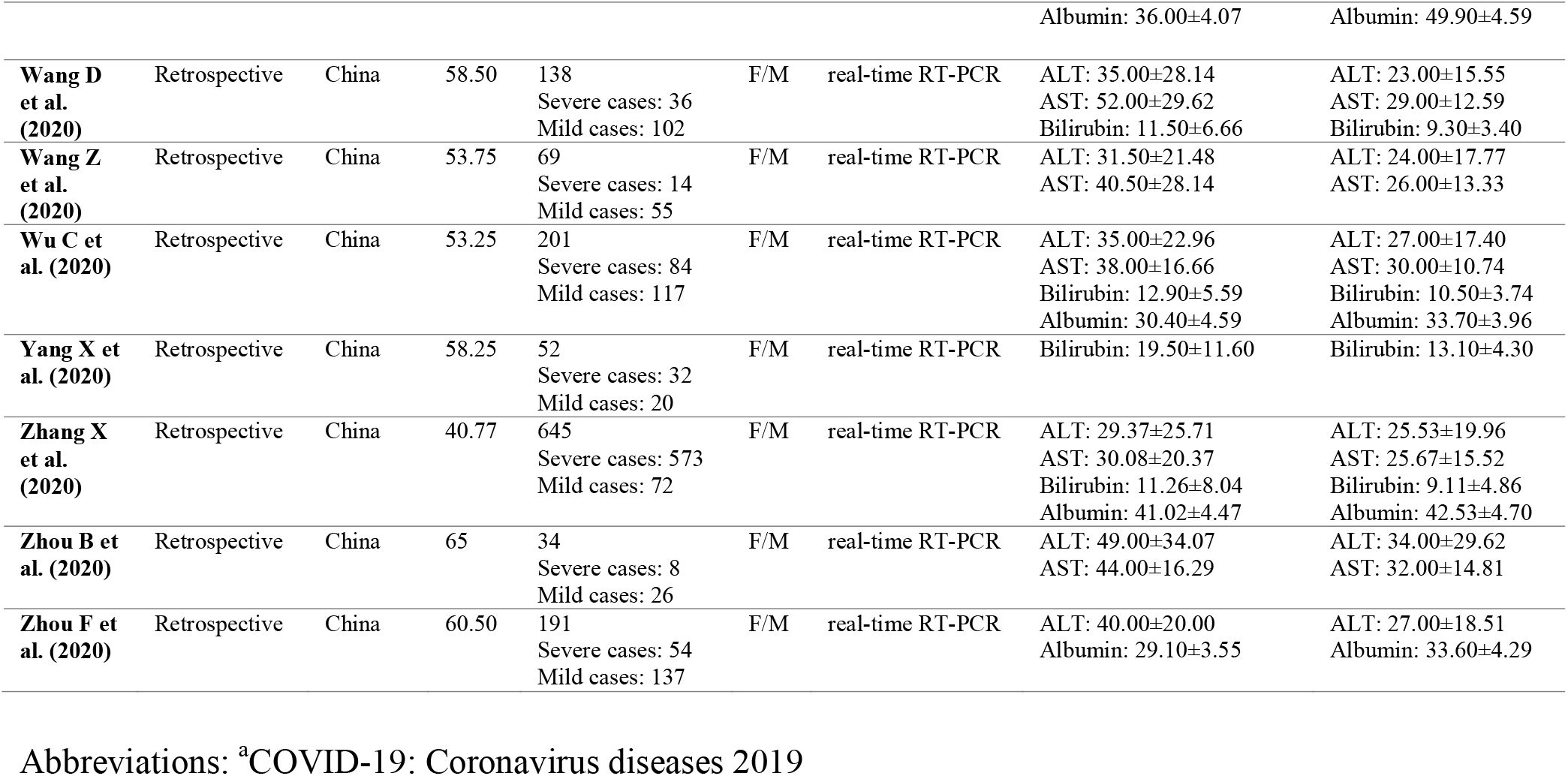
Characteristics of studies included in the meta-analysis.

### Serum levels of AST, ALT, total Bilirubin, Albumin and severity of COVID-19 infection

In the overall pooled estimate of 20 studies with 3,428 COVID-19 infected patients (severe cases = 1,455 and mild cases = 1,973), it was shown that higher serum levels of AST (weighted mean difference = 8.84 U/L, 95% CI = 5.97 to 11.71, P<0.001, I^2^ = 73.4%, P_heterogeneity_ <0.001) (Figure 2), ALT (weighted mean difference = 7.35 U/L, 95% CI = 4.77 to 9.93, P<0.001, I^2^ = 57.2%, P_heterogeneity_ = 0.001) (Figure 3) and total Bilirubin (weighted mean difference = 2.30 mmol/L, 95% CI = 1.24 to 3.36, P<0.001, I^2^ = 68.8%, P_heterogeneity_ <0.001) (Figure 4), were associated with a significant increase in the severity of COVID-19 infections. In addition, combined results from the random-effects model showed that lower serum levels of Albumin (weighted mean difference = −4.24 g/L, 95% CI = −6.20 to −2.28, P<0.001, I^2^ = 95.7%, P_heterogeneity_ <0.001) (Figure 5), significantly increased severity of the disease.

**Figure 2.**
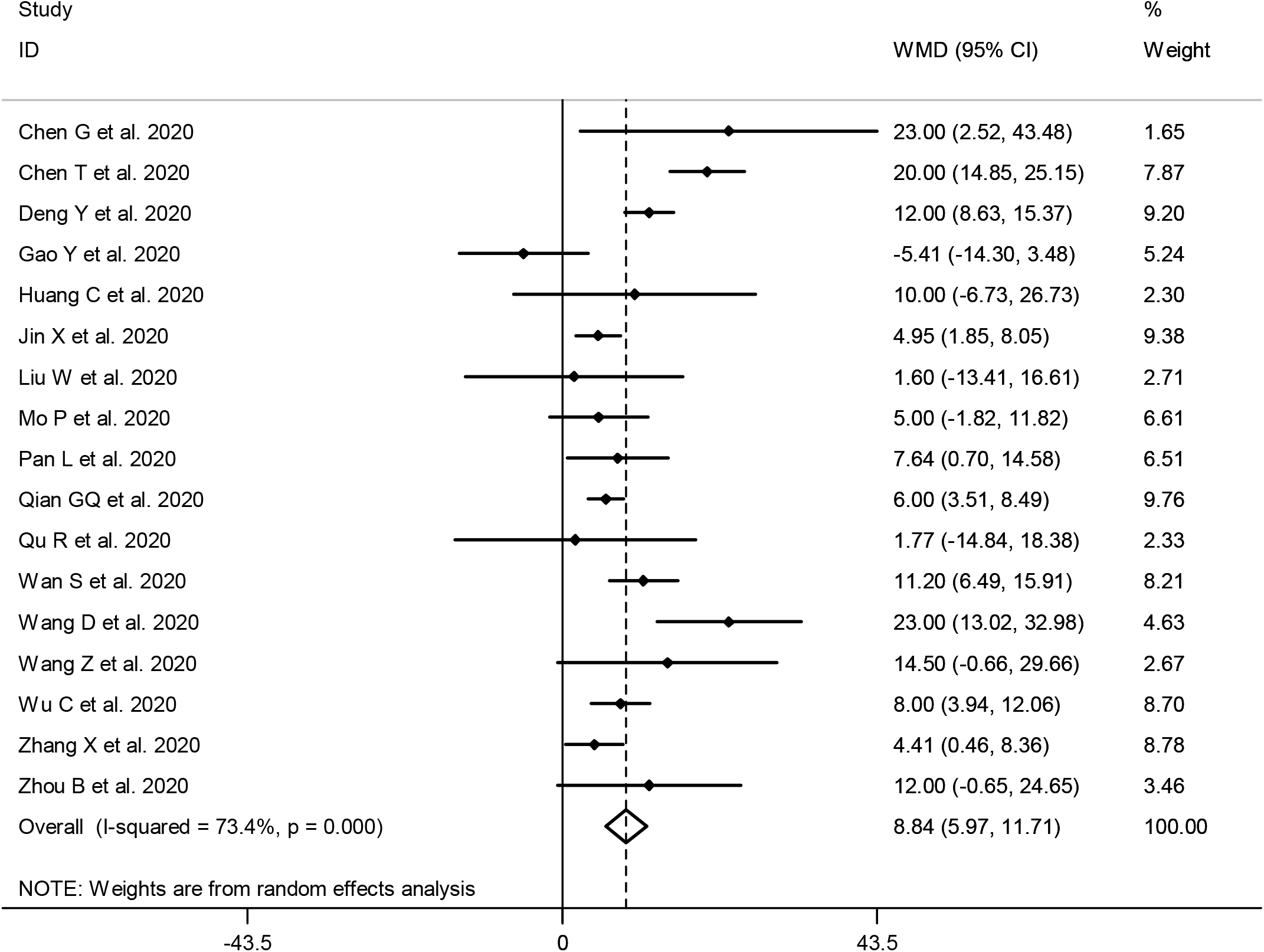
Forest plot for the association between serum levels of AST and severity of COVID-19 infection using random-effects model.

**Figure 3.**
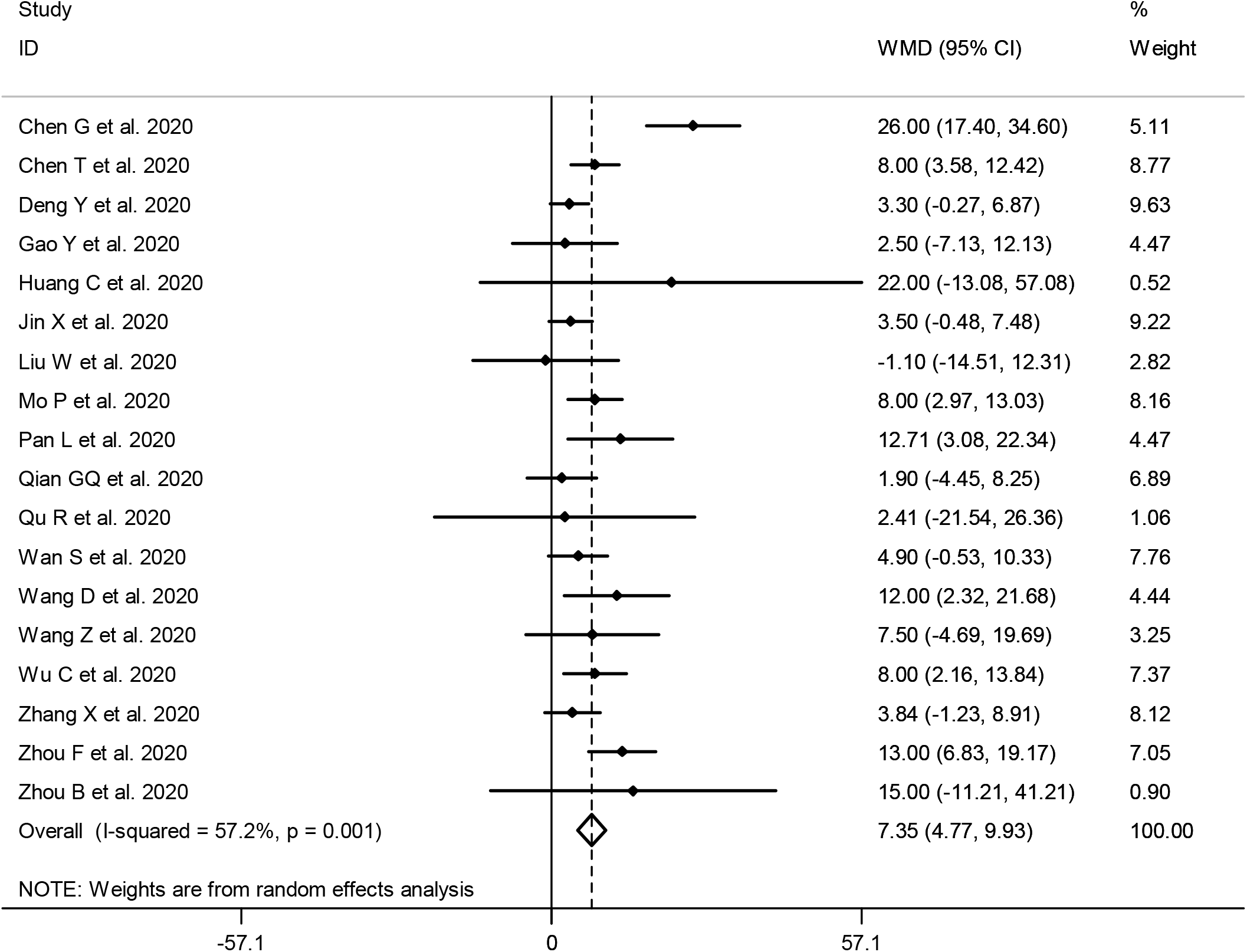
Forest plot for the association between serum levels of ALT and severity of COVID-19 infection using random-effects model.

**Figure 4.**
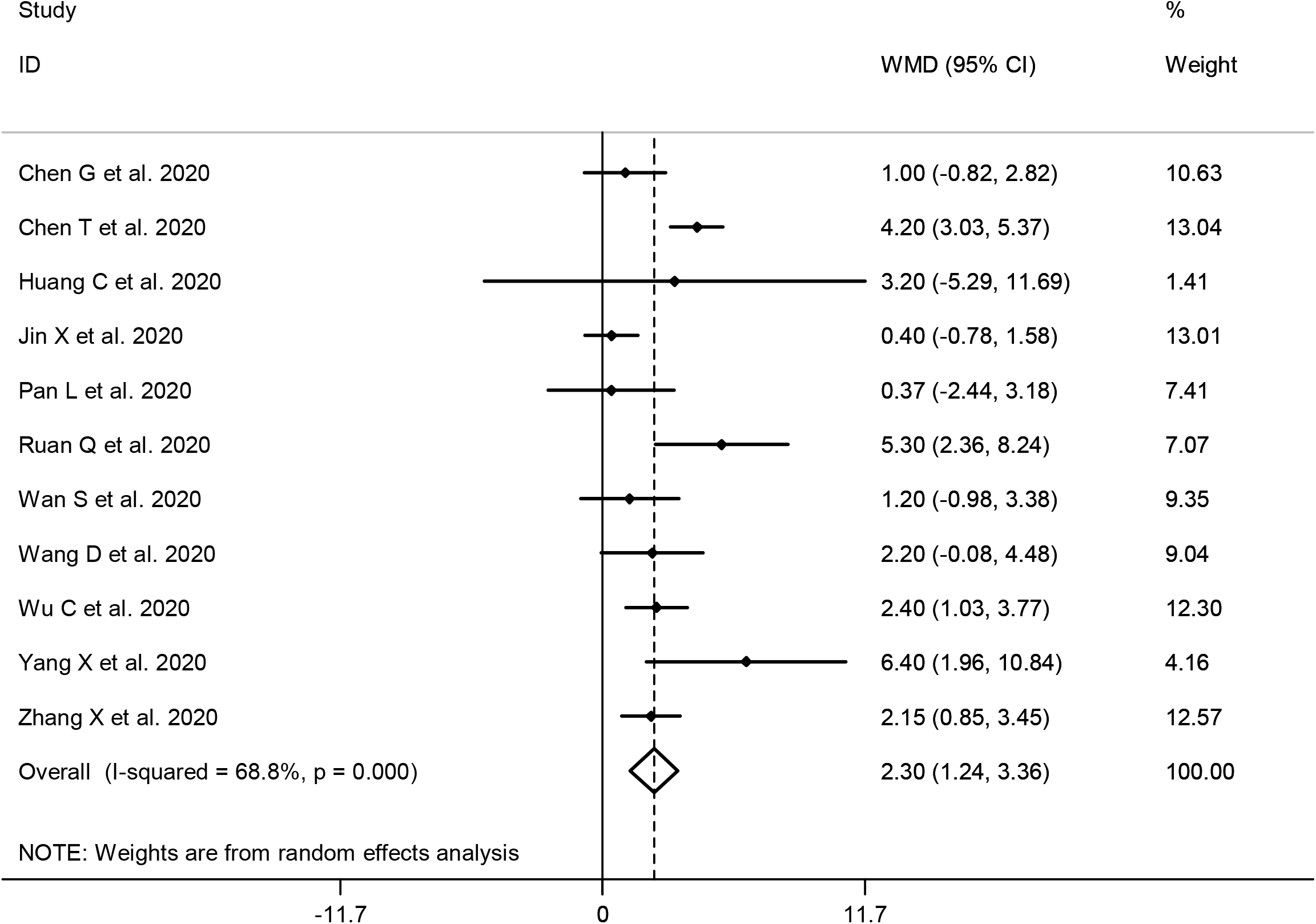
Forest plot for the association between serum levels of total Bilirubin and severity of COVID-19 infection using random-effects model.

**Figure 5.**
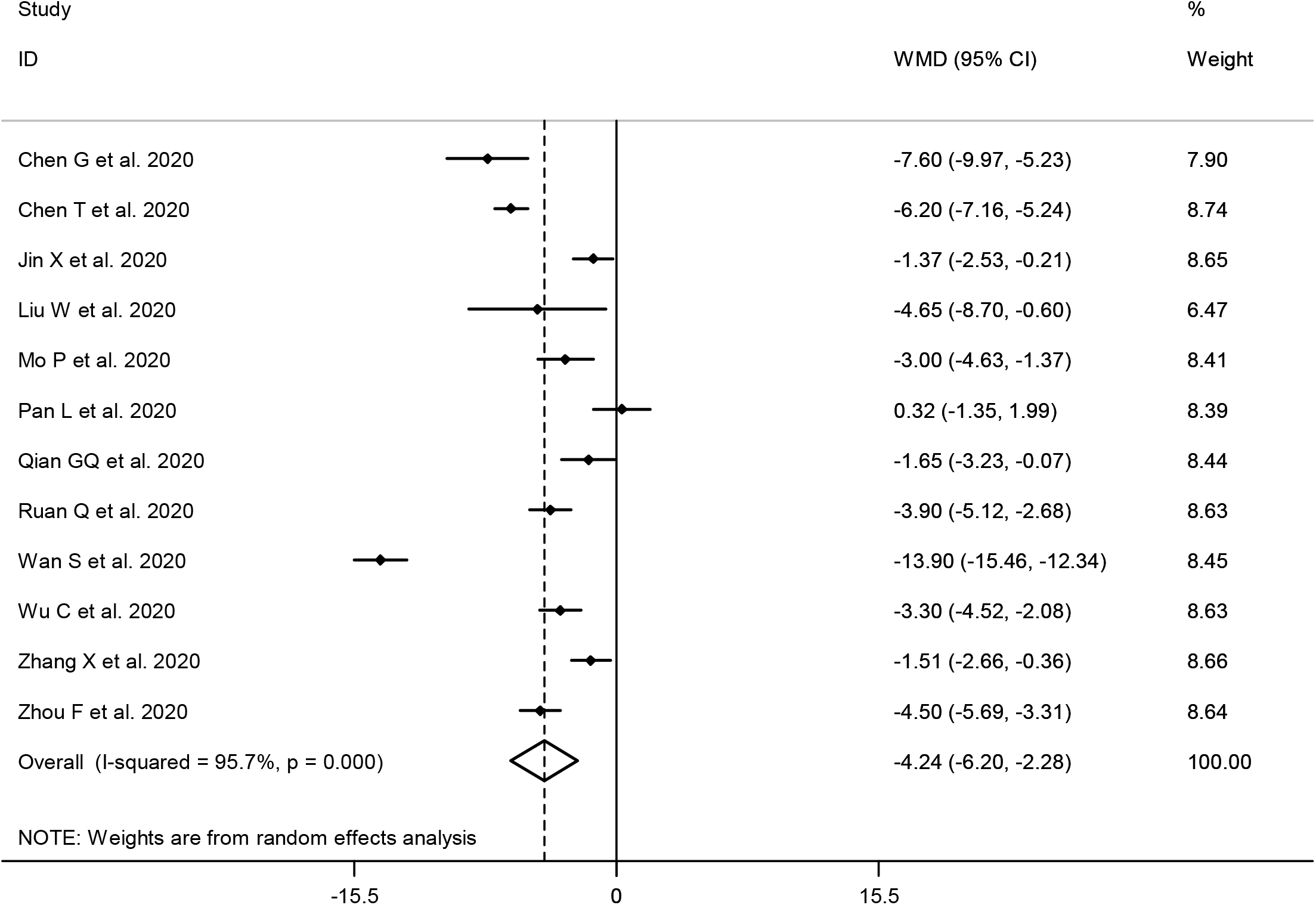
Forest plot for the association between serum levels of Albumin and severity of COVID-19 infection using random-effects model.

### Publication bias and sensitivity analysis

Based on the results of Egger’s test (AST; P = 0.465, ALT; P = 0.171, total Bilirubin; P = 0.663 and Albumin; P=0.802) and visual inspection of funnel plots, we found no evidence of publication bias (Supplementary Figures 1-4). Furthermore, findings from sensitivity analysis showed that overall estimates did not depend on a single study (Supplementary Figures 5-8).

## Discussion

Findings from this meta-analysis supported the hypothesis that liver injury is associated with severe outcomes in patients with COVID-19 infection. To our knowledge, this study is the first systematic review and meta-analysis to assess the association between serum levels of AST, ALT, total Bilirubin and Albumin with severity of COVID-19 infection.

Our results are in agreement with previous narrative review ^32^. Previously, liver damage has been reported as an important risk factor for severe outcome and death in SARS and MERS ^33-36^.

Mild cases of COVID-19 showed symptoms of dry cough, fever, fatigue, myalgia and diarrhea. In severe cases, viral pneumonia, dyspnea and hypoxemia occurred 1 week after the onset of the disease, which may progress to acute respiratory distress syndrome, metabolic acidosis, septic shock and even death ^12^. Previous studies have shown that the incidence of liver injury in severe COVID-19 patients ranged from 58% to 78% ^37, 38^, mainly indicated by elevated AST, ALT and total Bilirubin levels accompanied by slightly decreased Albumin levels ^12, 21, 24, 39^. Currently, studies on the mechanisms of COVID-19 related liver dysfunction are limited. COVID-19 uses the angiotensin converting enzyme 2 (ACE2) as the binding site to enter the host cell in lungs, kidneys and heart ^40^. Previous study showed that both liver and bile duct cells express ACE2 ^41^. In addition, the ACE2 expression of bile duct cells is much greater than that of liver cells. Bile duct epithelial cells are known to play important roles in initiation and regulation of immune responses and liver regeneration ^42^. So there is at least theoretical potential possibility of direct liver and bile duct involvement by the virus.

Serum concentrations of pro-inflammatory cytokines, including IL-1β, IL-6 and TNF-α increased in the majority of severe cases, suggesting cytokine storm syndrome might be associated with disease severity ^43^. Similarly, SARS and MERS were also characterized by exuberant inflammatory responses and end-organ damage ^44, 45^. Besides, the cytokine storm syndrome was observed in severe COVID-19 cases ^43^, yet whether it results in liver injury in patients remains to be investigated.

Mild lobular and portal activity along with moderate microvascular steatosis were observed in liver biopsy specimens, which might be caused by either COVID-19 infection or drug-induced liver injury ^46^. Similar to the situation in SARS and MERS, steroids, antivirals and antibiotics are widely used for the treatment of COVID-19 patients ^47-49^. Although these drugs are potential causes of liver dysfunction, there is little evidence that currently available drug combinations impair liver function in patients with COVID-19 infection ^24^. Actually, a recent study showed that the liver dysfunction might be caused by lopinavir/litonavir, which is used as antivirals for the treatment of COVID-19 patients ^50^.

The present study has some limitations. First, interpretation of our meta-analysis findings might be limited by the small sample size. Second, there is a lack of reports that liver failure occurs in COVID-19 patients with chronic liver diseases and our meta-analysis did not include data such as chronic hepatitis B or C.

## Conclusion

In this meta-analysis of 3,428 patients with confirmed COVID-19 in China, liver dysfunction was associated with severe outcome from COVID-19 infection. From a clinical perspective, attention should be paid to monitor the occurrence of liver dysfunction, and to the application of drugs which may induce liver injury, such as steroids and antibiotics of quinolone or macrolides. Patients with liver dysfunction are advised to be treated with drugs that could both inhibit inflammatory responses and protect liver functions, such as ammonium glycyrrhizinate ^51^, which in turn accelerates the process of disease recovery.

## Data Availability

All data are publicly available.

